# Endoscopic coagulation with clipping: a novel, simple, and effective strategy for colonic diverticular bleeding

**DOI:** 10.1101/2025.09.16.25335547

**Authors:** Toru Matsui, Chihiro Tanaka, Atsushi Horie, Yusuke Niwa, Masatoshi Satoh, Oki Nakano, Hiromitsu Oka, Yutaka Honda, Masaaki Takamura

**Author notes:** Corresponding author: Toru Matsui, MD, Department of Gastroenterology, Nagaoka Chuo General Hospital, 2041 Kawasaki-cho, Nagaoka City, Niigata 940-8653, Japan.

## Abstract

**Background and study aims:** Colonic diverticular bleeding (CDB) is a common cause of lower gastrointestinal bleeding, and rebleeding remains a significant clinical challenge despite various hemostatic techniques. Clipping and endoscopic band ligation are commonly performed; however, the efficacy of endoscopic coagulation with clipping (ECC) remains unclear. This study aimed to evaluate the efficacy and safety of ECC in treating CDB.

**Patients and methods:** We retrospectively analyzed 217 patients with suspected CDB seen at our institution between January 2017 and December 2024. Clinical outcomes, such as rebleeding rates within 30 days, adverse events, and length of hospital stay were assessed. ECC was defined as thermal coagulation of the bleeding vessel within the diverticulum, followed by closure with clips to reinforce hemostasis and prevent perforation.

**Results:** Of these, 202 patients underwent colonoscopic evaluation (15 were excluded due to spontaneous hemostasis or other reasons before endoscopy), 74 (36.6%) underwent ECC for hemostasis. Rebleeding occurred in 7 patients (9.5%) within 30 days. Diverticulitis with subsequent colonic perforation was observed in one patient (1.4%), who improved with conservative treatment, and exhibited no fatal adverse events.

**Conclusions:** With a favorable safety profile and low rebleeding rates, ECC can be a treatment option for CDB. Its simplicity and low reliance on specialized equipment make it a practical alternative to other hemostatic methods in clinical practice.

## Introductions

Colonic diverticular bleeding (CDB) is a common cause of acute lower gastrointestinal bleeding. Recent epidemiological studies in developed countries have indicated that the prevalence of colonic diverticulosis exceeds 50% in individuals aged over 60 years. Among these patients, CDB accounts for approximately 20% of acute lower gastrointestinal bleeding cases requiring hospitalization[1–3]. These conditions pose a significant burden on healthcare due to frequent rebleeding and readmission, with reported readmission rates ranging from 10%–20%[4]. Although mortality is generally low, it is significantly higher among older patients with comorbidities. These factors highlight the importance of effective hemostatic strategies in reducing clinical and economic burdens. Identifying the bleeding site in acute CDB remains challenging, with reported detection rates ranging from 17%–50%, depending on the timing and diagnostic modalities[5–8]. Inadequate localization of the bleeding site may increase the risk of rebleeding and necessitate repeat procedures. Therefore, accurate identification of the bleeding site and effective hemostasis are crucial for preventing recurrence.

Key strategies to improve identification have been proposed, including adequate bowel preparation, contrast-enhanced computed tomography (CT) before endoscopy[9], and emergency endoscopy within 24 h of onset[6, 8, 10, 11].

Once the source is identified, reliable hemostasis is required. Conventional endoscopic clipping is widely used; however, recent studies have reported superior outcomes with endoscopic band ligation (EBL). Reported rebleeding rates range from 9%–15% for EBL compared with 15–22% for clipping[2, 12–14]. However, EBL requires reinsertion of the endoscope and specialized equipment, which may limit its use in emergency settings. Other techniques, including over-the-scope clips (OTSC) and hemostatic powders like PuraStat, have been introduced but are limited by cost and uncertain efficacy[15, 16].

Endoscopic coagulation with clipping (ECC) is a technique involving thermal coagulation of the bleeding vessel followed by clip reinforcement, and it has been applied at our center as a practical and effective method for diverticular bleeding. Although coagulation therapy is the standard treatment for upper gastrointestinal bleeding, coagulation hemostasis is not considered the standard treatment for colonic bleeding worldwide. In institutions with experts in colorectal endoscopic submucosal dissection (ESD), thermal coagulation is commonly used for intraoperative hemostasis. For large lesions, prophylactic clip closure of the resection site is also performed to prevent delayed bleeding and perforation[17, 18]. This rationale also underlies ECC, in which post-coagulation clipping may reduce the risk of perforation and rebleeding. Nevertheless, coagulation hemostasis is generally avoided in CDB because the thin wall of the pseudodiverticulum increases the risk of perforation, and the safety of this approach remains under-investigated, with only a few reports evaluating adverse events. We therefore aimed to evaluate the efficacy and safety of ECC for the management of CDB.

## MATERIALS and METHODS

### Patients and Study Design

This study included patients with suspected CDB who underwent emergency colonoscopic evaluation at our institution between January 1, 2017, and December 31, 2024. A total of 217 patients were initially identified based on clinical presentation, such as hematochezia, and imaging findings consistent with diverticular bleeding. Among these, 202 patients underwent colonoscopy, 74 patients underwent ECC, and the remaining 128 were assigned to the non-ECC group, comprising 11 patients treated with endoscopic clipping alone and 115 who did not undergo any endoscopic hemostasis due to spontaneous cessation of bleeding or failure to localize the bleeding source, and 2 patients who underwent coagulation therapy alone without clip reinforcement. Patients were categorized according to the first therapeutic endoscopic procedure performed, regardless of whether the procedure was conducted during the initial or subsequent colonoscopy. For patients who initially showed spontaneous hemostasis but later required intervention due to rebleeding, the classification was based on the first active hemostatic method. The patient selection process and treatment categorization are summarized in Figure 1.

**Figure 1.**
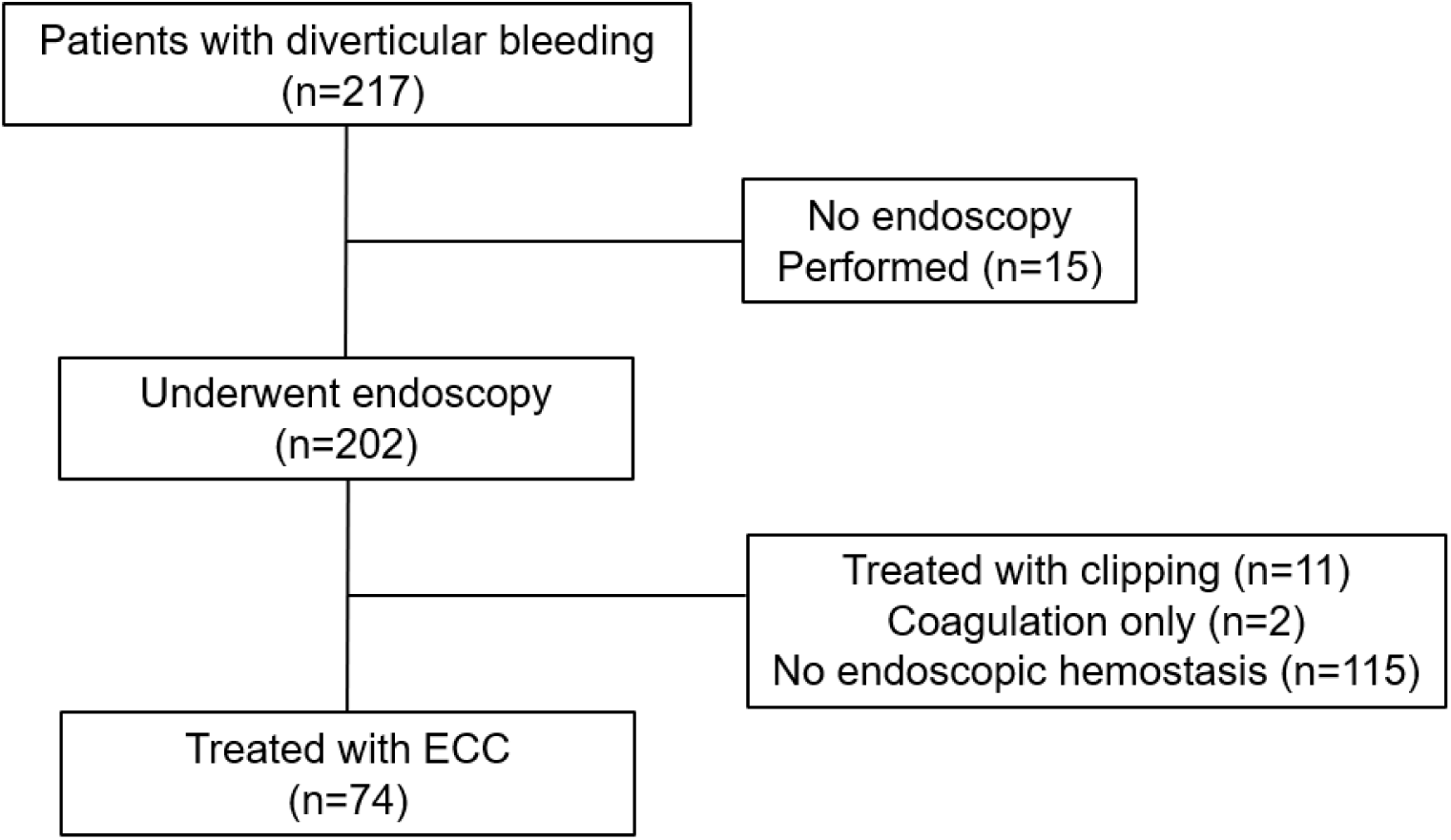
Flowchart of patient selection and treatment allocation. Of the 217 patients with suspected colonic diverticular bleeding, 202 underwent colonoscopy. Among these, 74 were treated with ECC, 11 with clipping alone, and 2 with coagulation alone (excluded from further analysis). The remaining 115 received no endoscopic hemostasis. ECC: endoscopic coagulation with clipping; CDB: colonic diverticular bleeding.

The study protocol was approved by the Institutional Review Board of Nagaoka Chuo General Hospital (reference number 688). All research methods were performed following the relevant guidelines and regulations.

### Inclusion and Exclusion Criteria

Patients were eligible if they were admitted with suspected CDB between January 2017 and December 2024 and had undergone emergency colonoscopy. CDB was suspected based on hematochezia and either (1) endoscopic confirmation of active bleeding or stigmata of recent hemorrhage (SRH) from a diverticulum or (2) CT evidence of extravasation in a patient with multiple colonic diverticula.

Exclusion criteria were (1) upper gastrointestinal bleeding; (2) other colonic bleeding sources such as tumors, ischemic colitis, or hemorrhoids; and (3) incomplete clinical records or transfer before undergoing colonoscopy.

### Definition of Terms

ECC was defined as thermal coagulation directly applied to the bleeding diverticulum, followed by clipping at the same site to reinforce hemostasis and prevent perforation. Early rebleeding was defined as rebleeding occurring within 30 days of initial hemostasis, as the responsible diverticulum could often be identified based on prior endoscopic findings. After this period, it becomes more difficult to identify the same lesion. In this study, “endoscopy within 24 h” referred to colonoscopy performed within 24 h after the patient’s arrival at the hospital—not from the onset of bleeding. Adverse events were defined as perforation, delayed bleeding, or clinically significant abdominal symptoms requiring additional management, assessed within 30 days post-procedure by review of medical records, imaging, and laboratory results.

### Endoscopic Procedures

Hemostasis was performed using a Coagrasper (FD-411UR; Olympus, Tokyo, Japan) with a high-frequency generator (ICC 200, VIO 300D; Erbe, Tübingen, Germany), and soft coagulation mode (Effect 4, 60 W) was used in most cases. The bleeding point was grasped with the forceps, gently lifted, and coagulated for 2–3 s (Figure 2). After coagulation, 2–4 standard hemostatic clips were typically applied to the treated diverticulum. The clips were placed in a manner that approximated the edges of the diverticular orifice, effectively closing it in a purse-string-like fashion to reinforce hemostasis and minimize the risk of delayed perforation. Representative procedures were recorded and are available as supplementary video materials.

**Figure 2.**
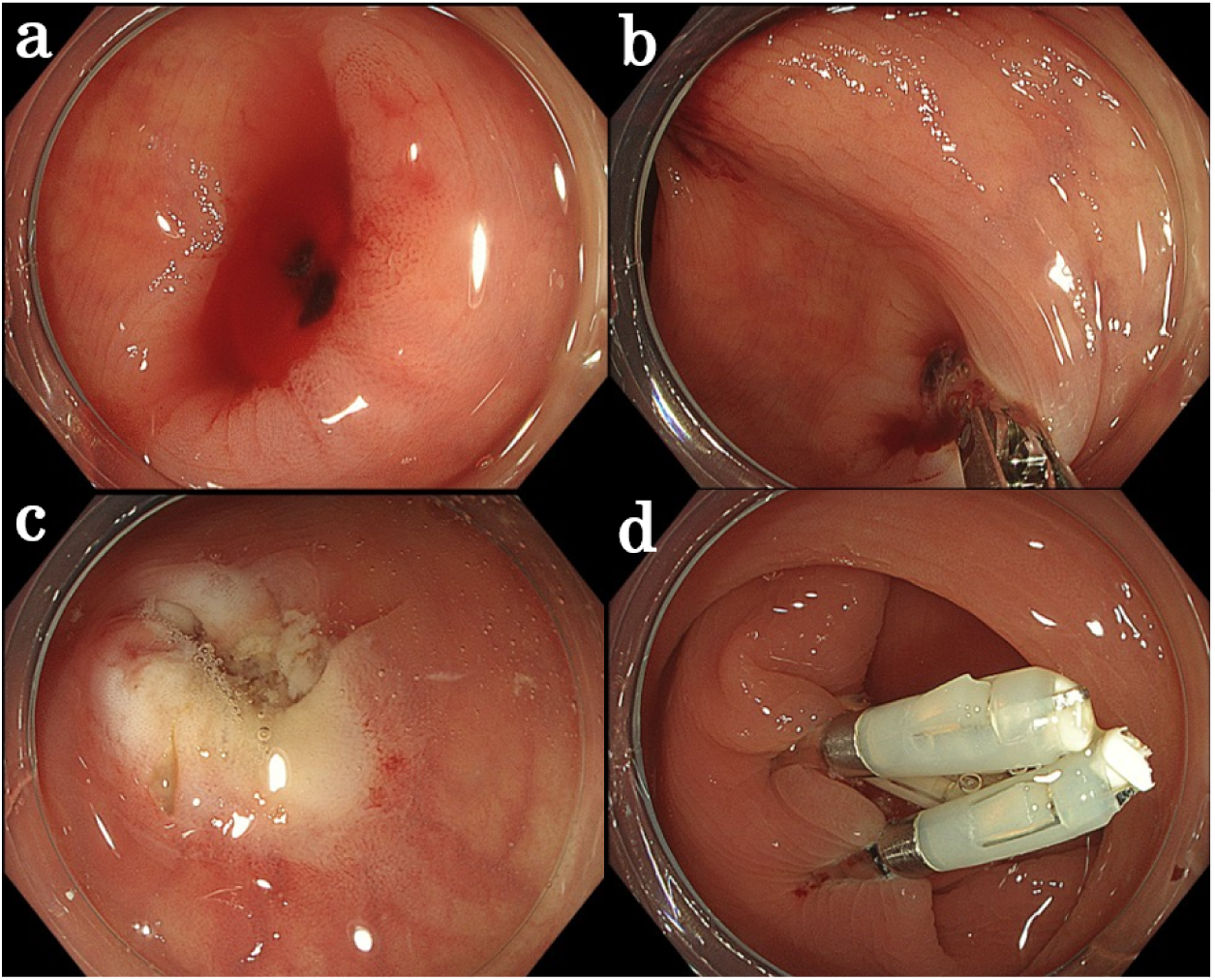
Hemostatic procedure using ECC. (a) A diverticulum identified as the bleeding source. (b, c) The bleeding site was grasped with monopolar hemostatic forceps, gently lifted, and coagulated. (d) The site was subsequently closed with clips to reinforce hemostasis and prevent perforation. ECC: endoscopic coagulation with clipping. **Supplementary Video 1**. Emergency colonoscopy revealed an actively bleeding diverticulum. Endoscopic coagulation with clipping (ECC) was successfully performed, resulting in immediate hemostasis without adverse events.

### Statistical Analysis

Statistical analyses were performed using EZR (Saitama Medical Center, Jichi Medical University), a graphical user interface for R with additional functions for biostatistics. Continuous variables were compared using the Mann–Whitney U test, and categorical variables were analyzed using Fisher’s exact test.

Statistical significance was defined as a 2-sided *P* < 0.05.

Exploratory univariate analysis was conducted to identify potential clinical factors associated with early rebleeding within 30 days in the ECC group. These analyses were considered hypothesis-generating and interpreted with caution due to the small number of rebleeding cases (n = 7).

## RESULTS

Table 1 summarizes the baseline characteristics of patients in the ECC and non-ECC groups. The ECC group had a significantly higher proportion of patients with a history of cerebral infarction (18.9% vs. 7.8%, *P* = 0.024) and showed a trend toward more frequent use of NSAIDs (18.9% vs. 9.4%, *P* = 0.079). The frequency of antithrombotic agent use did not differ significantly between the groups (43.2% vs. 32.0%, *P* = 0.129). The incidence of contrast extravasation on CT was significantly higher in the ECC group (51.4% vs. 19.5%, *P* < 0.001), suggesting more active or localized bleeding in patients selected for ECC. Overall, baseline characteristics, including age, comorbidities, and medication use, were similar between the ECC and non-ECC groups, minimizing potential selection bias. In the ECC group, 7 of the 74 patients (9.5%) experienced rebleeding within 30 days, including 6 cases (8.1%) within the first week. Table 2 details their clinical courses, with 3 undergoing repeat ECC, 1 endoscopic clipping, and 1 transcatheter arterial embolization followed by surgery; 2 required no further intervention. In 2 of the repeat ECC cases, bleeding originated from a different diverticulum than the initial site.

**Table 1.** Baseline characteristics of patients in the ECC and non-ECC groups.

| Variable | ECC group<br>(n = 74) | Non-ECC group<br>(n = 128) | *P* value |
| --- | --- | --- | --- |
| Age, years, median [IQR] | 75.0 [67.5–82.8] | 78.0 [65.0–84.0] | 0.627 |
| Male, n (%) | 57 (77.0%) | 84 (65.6%) | 0.112 |
| BMI, kg/m<sup>2</sup>, median [IQR] | 23.2 [20.0–24.5] | 22.9 [20.4–24.7] | 0.113 |
| Diabetes, n (%) | 21 (28.4%) | 29 (22.7%) | 0.400 |
| Hypertension, n (%) | 51 (68.9%) | 74 (57.8%) | 0.134 |
| Renal dysfunction (eGFR < 60), n (%) | 34 (45.9%) | 44 (34.4%) | 0.133 |
| Ischemic heart disease, n (%) | 11 (14.9%) | 13 (10.2%) | 0.369 |
| Cerebral infarction, n (%) | 14 (18.9%) | 10 (7.8%) | 0.024 |
| NSAID use, n (%) | 14 (18.9%) | 12 (9.4%) | 0.079 |
| Antithrombotic use, n (%) | 32 (43.2%) | 41 (32.0%) | 0.129 |
| Rebleeding within 30 days, n (%) | 7 (9.5%) | 20 (15.6%) | 0.284 |
| Hospital stay, days, median [IQR] | 7.0 [5.0–9.8] | 7.0 [6.0–9.2] | 0.344 |
| Blood transfusion, n (%) | 22 (29.7%) | 38 (29.2%) | 1.000 |
| Hemoglobin before colonoscopy, g/dL,<br>median [IQR] | 9.8 [7.7–11.1] | 10.2 [8.3–11.6] | 0.329 |
| CT extravasation, n (%) | 38 (51.4%) | 25 (19.5%) | <0.001 |
| Colonoscopy within 24 h, n (%) | 66 (89.2%) | 79 (61.7%) | <0.001 |

Abbreviations: ECC, endoscopic coagulation with clipping; IQR, interquartile range; BMI, body mass index; eGFR, estimated glomerular filtration rate; NSAID, non-steroidal anti-inflammatory drugs; CT, computed tomography.

**Table 2.** Clinical outcomes in the ECC group.

| **Clinical outcome** | **ECC group**<br>**(n = 74)** | **Remarks** |
| --- | --- | --- |
| Rebleeding within 30 days | 7 / 74 (9.5%) | One case occurred 12 days after initial treatment. |
| Rebleeding within 7 days | 6 / 74 (8.1%) |  |
| **Interventions at rebleeding:** |  |  |
| – ECC retreatment | 3 cases | Two cases rebled from diverticula different from the initial treatment site. |
| – Endoscopic clipping | 1 case |  |
| – TAE followed by surgery | 1 case | Endoscopic intervention was technically challenging due to poor scope maneuverability. |
| – Observation only | 2 cases |  |
| **Delayed perforation** | 1 case (1.4%) | Resolved with conservative management. |

Abbreviations: ECC, endoscopic coagulation with clipping; TAE, transcatheter arterial embolization.

One patient (1.4%) developed delayed perforation, which was managed conservatively, and no other severe adverse events were observed. Table 3 compares the clinical characteristics of the patients in the ECC group with and without rebleeding. Median hemoglobin levels and the frequency of blood transfusion were comparable between the rebleeding and non-rebleeding groups (8.8 vs. 10.0 g/dL, *P* = 0.554; 42.9% vs. 28.4%, *P* = 0.420). Emergency colonoscopy within 24 h was performed at similar rates in the non-rebleeding and rebleeding groups (89.6% vs. 85.7%, *P*=1.000). SRH were defined as endoscopic findings suggestive of recent bleeding activity, including AB, NBVV, or AC, similar to Forrest I or □ lesions in upper gastrointestinal hemorrhage. Interestingly, contrast extravasation was observed less frequently in patients who experienced rebleeding (28.6% vs. 53.7%, *P* = 0.255).

**Table 3.** Comparison of clinical characteristics between rebleeding and non-rebleeding groups in the ECC cohort. Variables selected based on clinical relevance to bleeding severity, treatment decision-making, or rebleeding risk.

| Clinical variable | Rebleeding group (n = 7) | Non-rebleeding group (n = 67) | P value |
| --- | --- | --- | --- |
| Age (years), median [IQR] | 72.0 [68.5–81.0] | 75.0 [68.0–82.5] | 0.919 |
| BMI (kg/m<sup>2</sup>), median [IQR] | 21.9 [20.2–24.7] | 23.4 [21.6–25.6] | 0.401 |
| Hemoglobin (g/dL), median [IQR] | 8.8 [8.3–9.4] | 10.0 [7.7–11.1] | 0.554 |
| Antithrombotic drug use, n/N (%) | 3/7 (42.9%) | 30/67 (44.8%) | 1.000 |
| CT extravasation, n/N (%) | 2/7 (28.6%) | 36/67 (53.7%) | 0.255 |
| SRH classification | AB: 3, AC: 4, NBVV: 0 | AB: 49, AC: 13, NBVV: 5 | - |
| Blood transfusion, n/N (%) | 3/7 (42.9%) | 19/67 (28.4%) | 0.418 |
| Endoscopy within 24 h, n/N (%) | 6/7 (85.7%) | 60/67 (89.6%) | 0.567 |

Abbreviations: ECC, endoscopic coagulation with clipping; IQR, interquartile range; SRH, stigmata of recent hemorrhage; AB, active bleeding; NBVV, non-bleeding visible vessel; AC, adherent clot.

As shown in Table 4, the rebleeding rate tended to be lower in patients with AB-type SRH (5.8%) than in those with AC or NBVV types (18.2%, *P* = 0.186), possibly due to better target visibility, allowing more precise hemostasis using ECC.

**Table 4.**
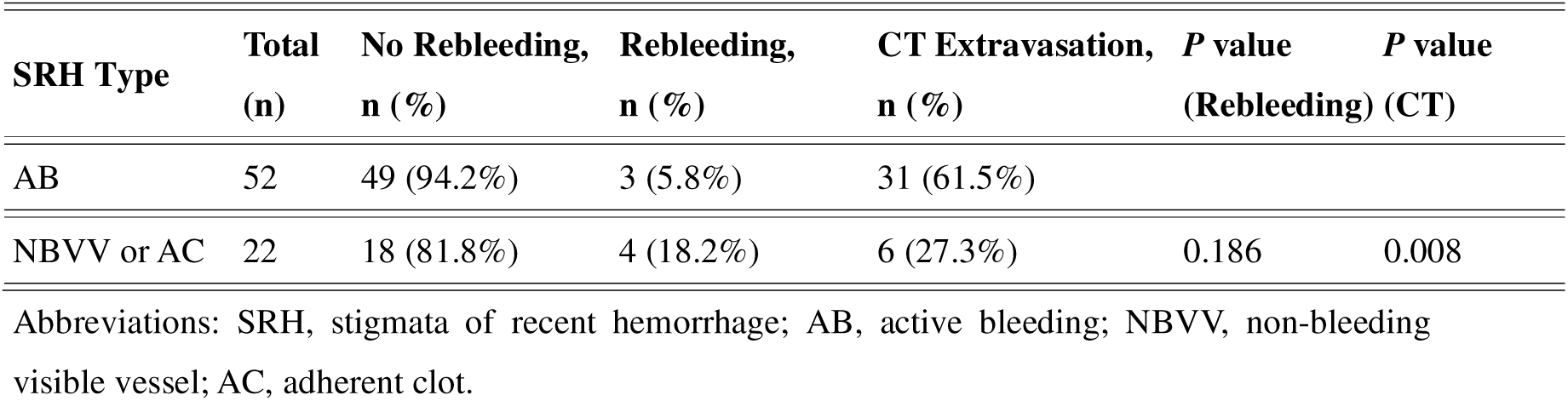
Comparison of the rebleeding rates and CT extravasation between patients with AB and NBVV/AC-type SRH.

## DISCUSSION

Because spontaneous hemostasis is relatively common in colonic diverticular bleeding, endoscopic intervention is generally reserved for cases with evident or strongly suspected bleeding. In particular, intervention is recommended when stigmata of recent hemorrhage (SRH)—such as AB, NBVV, or AC—are present [19–22]. Accordingly, ECC was mainly performed when the bleeding point was clearly identified and endoscopically accessible, reflecting real-world decision-making in acute lower gastrointestinal bleeding. This selection inevitably introduces bias, but it also allows that ECC is applied in scenarios where hemostasis is most likely to be technically feasible and effective. Endoscopic clipping remains the most common technique for CDB. Its effectiveness, however, has been increasingly questioned, particularly in cases where indirect clipping is performed without clear visualization of the bleeding source, which has been associated with relatively high rebleeding rates[23]. Recent studies have suggested that direct clipping, in which the bleeding vessel is directly targeted, results in significantly lower rebleeding rates than indirect clipping. In our study, 11 patients treated with endoscopic clipping were included in the non-ECC group; however, this subgroup was not analyzed separately due to the small sample size and the lack of detailed information regarding whether direct or indirect clipping was performed. This reflects a common limitation in retrospective studies, where distinguishing between the direct and indirect approaches is often difficult [24].

In our cohort, patients with AB-type SRH, representing active bleeding, had a markedly lower rebleeding rate than those with NBVV- or AC-type SRH (5.8% vs. 18.2%). This difference likely reflects the greater visibility and accessibility of the bleeding point in AB-type cases, facilitating more accurate and effective hemostasis using ECC. Although this difference did not reach statistical significance, the trend warrants confirmation in larger, multicenter studies. In contrast, NBVV- or AC-type lesions may involve either less clearly defined targets, making durable hemostasis more difficult to achieve. These results underscore the importance of SRH type in guiding both treatment decisions and prognostic expectations for diverticular bleeding, and also highlight the challenges in comparing outcomes between different hemostatic techniques, particularly in retrospective analysis.

Given these challenges, especially in cases with limited visibility such as indirect clipping or NBVV/AC-type SRH, EBL is gaining attention as a potentially superior option. Supported by accumulating evidence, it is increasingly reported as effective in selected cases with visualized SRH. Accordingly, many studies and guidelines now describe endoscopic clipping as “feasible but inferior to EBL,” and EBL has become increasingly popular[13, 25, 26]. However, despite its lower rebleeding rate in several studies, EBL has practical limitations. The technique requires withdrawal and reinsertion of the endoscope and the use of dedicated banding devices. Moreover, EBL is only applicable when a definite bleeding point is visualized, typically in patients with SRH. Consequently, its use may be constrained in real-world practice.

In contrast to other hemostatic approaches, the use of coagulation hemostasis in CDB warrants attention. Unlike in the upper gastrointestinal tract, coagulation is not frequently used in CDB, likely due to concerns about perforation. Anatomically, diverticula form at sites where blood vessels penetrate the muscularis propria, creating natural weaknesses in the intestinal wall[27]. Furthermore, thermal injury to thin-walled structures may increase the risk of adverse events, including delayed perforation.

This risk may be particularly relevant in older patients, in whom comorbidities and medications are also more prevalent. Conditions such as hypertension and diabetes, as well as the use of antiplatelet or anticoagulant agents can contribute to vascular fragility and reduced coagulation, thereby increasing the risk of rebleeding[28].

Against this backdrop, the risks associated with thermal injury merit careful consideration. Although adverse events related to coagulation therapy have been reported, such as post-polypectomy coagulation syndrome (PPCS) during polypectomy procedures[29, 30], there have been no comprehensive reports of serious adverse events such as perforation. The safety and efficacy of coagulation-based hemostatic techniques for CDB have likewise not yet been thoroughly evaluated. One factor that may influence the safety profile of coagulation therapy is the type of electrosurgical device used. In monopolar devices, electrical current travels from the active electrode through the patient’s body to the return electrode. In contrast, bipolar devices deliver current only through tissues located between the two electrodes of the instrument[31–33]. Although some institutions prefer bipolar devices to minimize the risk of deep-tissue injury, monopolar hemostasis using soft coagulation with low-power settings has been optimized to reduce thermal penetration.

ECC is usually performed by directly targeting the bleeding site; however, the confined space within a diverticulum can hinder precise grasping.

In such cases, thermal coagulation slightly adjacent to the presumed bleeding point often results in successful hemostasis. One case of delayed perforation occurred (1.4%); however, no immediate perforation or procedure-related deaths were observed. The adverse event rate observed is comparable to or even lower than the reported adverse events associated with polypectomy or mucosal resection. PPCS, previously described in association with thermal devices in other endoscopic procedures, was not observed in our series. However, in our ECC cohort, only 1 case of delayed perforation occurred, with no immediate serious adverse events, suggesting a favorable safety profile.

Importantly, ECC achieved a rebleeding rate of 9.5%, which was comparable to or better than the rates reported for EBL in previous studies[12, 13]. ECC is technically straightforward and cost-effective, without the need for specialized devices—unlike hemostatic powders (e.g., PuraStat) or OTSC systems, which may require additional equipment and cost[34, 35]. Because of its favorable safety profile, technical simplicity, and low rebleeding rate, ECC represents an effective hemostatic strategy that can be readily adopted in routine clinical settings. Our study has some limitations.

First, this was a retrospective, single-center study without a direct comparator group, and the observation period was limited to 30 days; thus, long-term outcomes remain unknown. Second, most ECC-treated diverticular bleeding cases occurred in the right colon (69%), reflecting the general distribution pattern observed in Asian populations. As left-sided bleeding is more common in Western countries, the applicability to those settings may be limited, although ECC was also successfully applied to left-sided cases in this study.

Third, there is a potential selection bias as ECC and EBL are generally performed only in patients with clearly identified SRH, whereas clipping is often used even when SRH is absent. This may partly account for the differences in rebleeding rates among techniques. While multivariate analysis was not feasible due to the limited number of rebleeding cases, the consistency of outcomes in this cohort suggests that ECC may still represent a practical and reproducible option for selected patients. Further multicenter studies are warranted to assess its applicability in broader clinical settings. In conclusion, ECC achieved a low rebleeding rate without the need for specialized devices and was technically simple to perform. These results indicate that ECC is a feasible option for routine clinical practice in managing colonic diverticular bleeding.

## Data Availability

All data generated or analyzed during this study are available from the corresponding author on reasonable request.

